# Metabolomic changes associated with frontotemporal lobar degeneration syndromes

**DOI:** 10.1101/2020.02.10.20021758

**Authors:** Alexander G. Murley, P Simon Jones, Ian Coyle Gilchrist, Lucy Bowns, Julie Wiggins, Kamen A. Tsvetanov, James B. Rowe

**Author notes:** Corresponding Author Alexander Murley, University of Cambridge,.

## Abstract

**Objective:** Widespread metabolic changes are seen in neurodegenerative disease and could be used as biomarkers for diagnosis and disease monitoring. They may also reveal disease mechanisms that could be a target for therapy. In this study we looked for blood-based biomarkers in syndromes associated with frontotemporal lobar degeneration.

**Methods:** Plasma metabolomic profiles were measured from 134 patients with frontotemporal lobar degeneration (behavioural variant frontotemporal dementia n=30, non fluent variant primary progressive aphasia n=26, progressive supranuclear palsy n=45, corticobasal syndrome n=33) and 32 healthy controls.

**Results:** Forty-nine of 842 metabolites were significantly altered in frontotemporal lobar degeneration (after false-discovery rate correction for multiple comparisons). These were distributed across a wide range of metabolic pathways including amino acids, energy and carbohydrate, cofactor and vitamin, lipid and nucleotide pathways. The metabolomic profile supported classification between frontotemporal lobar degeneration and controls with high accuracy (88.1-96.6%) while classification accuracy was lower between the frontotemporal lobar degeneration syndromes (72.1-83.3%). One metabolic profile, comprising a range of different pathways, was consistently identified as a feature of each disease versus controls: the degree to which a patient expressed this metabolomic profile was associated with their subsequent survival (hazard ratio 0.74 [0.59-0.93], p = 0.0018).

**Conclusions:** The metabolic changes in FTLD are promising diagnostic and prognostic biomarkers. Further work is required to replicate these findings, examine longitudinal change, and test their utility in differentiating between FTLD syndromes that are pathologically distinct but phenotypically similar.

## Introduction

Frontotemporal lobar degeneration (FTLD) causes a wide spectrum of syndromes including the behavioural and language variants of frontotemporal dementia (bvFTD, PPA respectively), progressive supranuclear palsy (PSP) and corticobasal syndrome (CBS).[1, 2] Accurate early diagnosis is challenging, due in part to the specialist clinical skills and imaging resources required. There is therefore a pressing need for FTLD biomarkers. Such biomarkers may also facilitate clinical trials monitoring and reveal disease mechanisms as a target for therapy. In this study we looked for blood-based metabolic biomarkers in four clinical syndromes associated with FTLD. We studied the four syndromes together, in view of their potential commonalities in clinical and neuropathological features.[1, 3, 4]

Metabolic pathways are likely to be altered in FTLD. For example, genomic studies of FTLD syndromes have identified gene loci polymorphisms implicated in a range of metabolic processes including protein synthesis, packaging and breakdown, as well as immune functions and myelin structure.[5–9] In addition, the identification, quantification and analysis of metabolic pathways using metabolomics has identified candidate biomarkers in other neurodegenerative diseases including Alzheimer’s, Huntington’s and Parkinson’s diseases.[10–12] However, there is limited evidence on metabolomic abnormalities in FTLD: the cerebrospinal fluid in FTD shows a panel of metabolites could differentiate FTD from controls and Alzheimer’s disease,[13] while hypertriglyceridemia and hypoalphalipoproteiemia have been reported in bvFTD.[14]

This study had three aims. First, to identify which biochemicals and their associated metabolite pathways are abnormal in each of four FTLD syndromes. Second, to test the accuracy of metabolite profiles in classifying patients versus healthy controls. Third, to test whether metabolomics changes are indicative of prognosis. We predicted that a wide range of metabolic pathways would be abnormal in FTLD and it would be possible to accurately classify between FTLD syndromes and controls; but that phenotypic and pathological similarities would reduce the accuracy of differential diagnosis between the FTLD syndromes.

## Materials and Methods

### Study Participants

Patients were recruited from the Cambridge Centre for Frontotemporal Dementia and Related Disorders and met the clinical diagnostic criteria for either behavioural variant frontotemporal dementia[15], non-fluent variant primary progressive aphasia[16], progressive supranuclear palsy Richardson’s syndrome[17] or corticobasal syndrome.[18] Healthy controls had no neurological or psychiatric disease. The study was approved by the local ethics committee and all participants gave informed consent or, if lacking mental capacity, through a consultee process according to UK law. 134 patients (30 bvFTD, 26 nfvPPA, 45 PSP, 33 CBS) and 32 healthy controls participated. Plasma was obtained by centrifugation of whole blood and stored at -80°C until analysis.

### Metabolite detection and quantification

Biochemical identification and quantification was performed by Metabolon Inc (www.metabolon.com) for all samples at a single timepoint. Samples were analysed with ultra-high performance liquid chromatography and tandem mass spectrometry, optimised for basic and acidic species. Biochemicals were then identified by comparison of the ion features of each sample to a reference library of compounds and grouped into sub and super pathways, corresponding to metabolite pathways[19]. For a full list of the metabolic pathways and their constituent biochemicals measured in this study see Appendix 1.

### Statistical analysis

Our statistical analysis pipeline is summarised in Figure 1. First, we used independent two-sample *t-*tests to compare the age distributions of the FTLD and control groups. A Chi-squared test with Yates correction was used to compare sex between groups. In the metabolite dataset missing values implied a result below the limit of detection in that individual. We excluded metabolites if they were missing in more than half of the participants. Remaining missing values were replaced by half of the minimum positive value of that variable. We also removed metabolites from exogenous metabolic pathways, including known drugs and drug pathways, before further analysis. All metabolite concentrations were scaled to unit variance (i.e. normalised to z-scores). [20]

**Figure 1:**
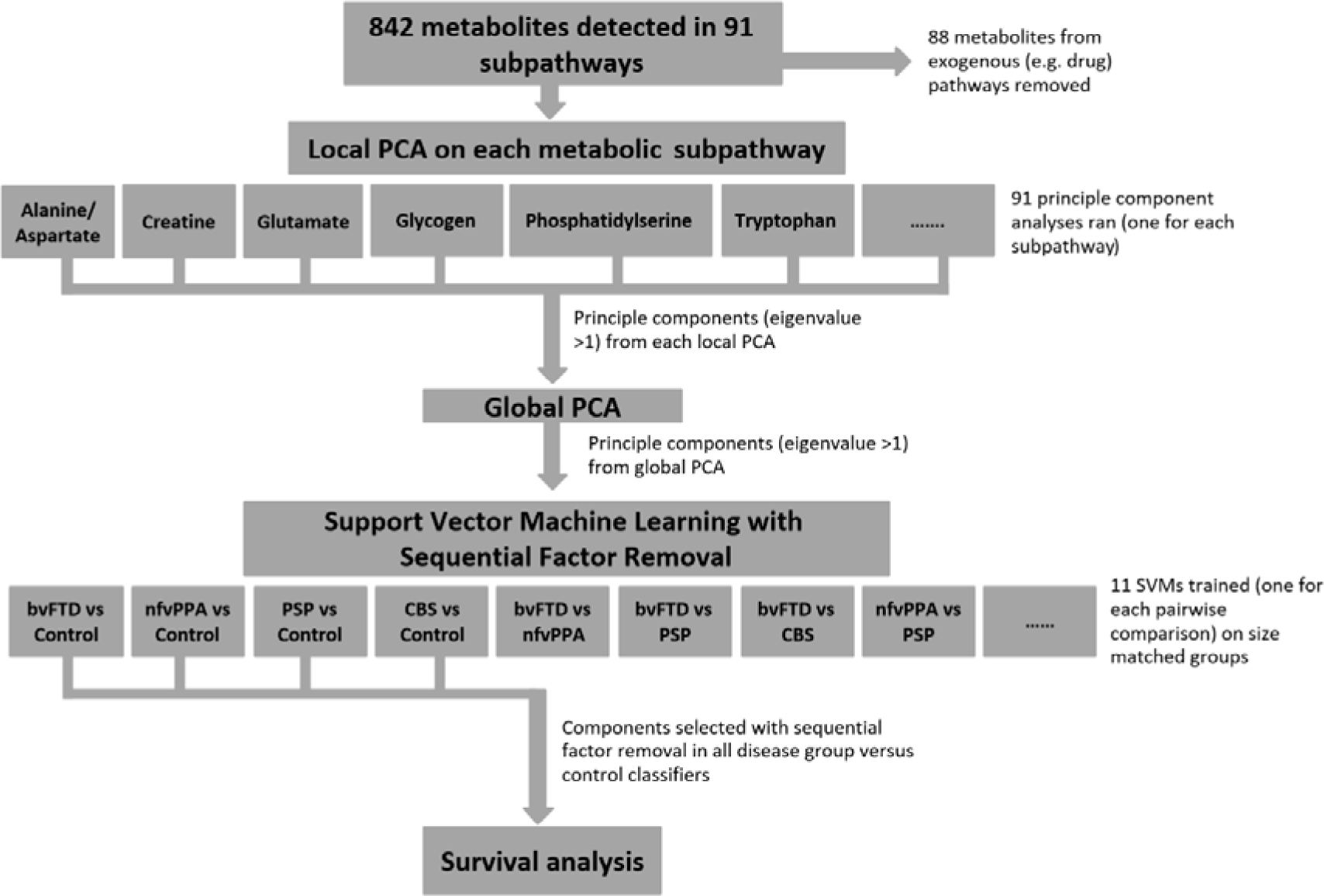
Summary of the analysis pipeline. From a total of 842 metabolites, a principal component analysis (PCA) was run on the metabolites in each of 91 subpathways. All components with eigenvalue greater than 1 were entered into a global PCA. The subject-specific weights of the principal components from this PCA were used as features for support vector machines, using k-fold cross-validation and recursive feature elimination. Components selected by recursive feature elimination were then used as predictors for the survival analysis (cox proportional hazards regression with age, gender and FTLD subgroup as covariates).

Univariate statistical tests were then used to compare individual metabolite differences between groups. We used a generalised linear model on each metabolite, with age and sex as covariates, to compare the FTLD and control groups. An FDR corrected p-value threshold of 0.01 used to determine statistical significance (using the ‘mafdr’ function in MATLAB). Bonferroni correction is also presented, while noting that non-independence of metabolites is likely to make this method overly conservative. Fold change for each metabolite was calculated by dividing the mean disease and control values of unscaled data.

A two-level principal component analysis (PCA) was used to explore the diseases’ effect on each metabolite pathway. We used this two-level approach to reduce dimensionality whilst preserving the metabolite pathways structure of the dataset, with parsimonious representation of all metabolic pathways in the comparisons between groups. At the first level, we performed a ‘local PCA’ on the metabolites in each sub pathway, to identify components that best explained the variance in that pathway. Ninety-one local principle component analyses were run in total, one for each metabolite subpathway. Within each subpathway, we used the Kaiser criteria to select components with an eigenvalue greater than one. To assess which metabolite pathways are affected in FTLD we used independent two-sample *t*-tests to compare scores for each local PCA component. An FDR corrected p-value threshold of 0.01 was used to determine statistical significance. At the second level, we performed a ‘global PCA’. This was global in the sense of examining metabolite variance across all subpathways, including all the components with an eigenvalue greater than one from all local PCAs.

Next we tested the ability of the global PCA components to classify FTLD syndromes. We trained pairwise linear support vector machines using the subject specific weightings for components output from the global PCA. A total of eleven SVMs were trained, to discriminate between each of the five groups, and to compare all FTLD syndromes *jointly* versus healthy controls. Prior to training, component loading values were rescaled from -1 to 1. Groups were size-matched by randomly sampling cases from the larger of the groups to match the size of the smaller group.

We used backwards sequential feature selection using the ‘sequentialfs’ function in MATLAB to identify the components that best predicted disease, as follows. Starting with the full dataset, components were sequentially removed until classifier accuracy decreased. SVM accuracy and factor selection was validated with 10-fold cross validation. In each iteration, the training and test data subsets were kept separate. Random case sampling, SVM training and sequential feature selection was repeated 10 times and the mean accuracy over all partitions was calculated. Only the components selected in all repetitions are reported. With small sample sizes, k-fold cross-validation minimises the bias of within-sample cross-validation.[21] The reported accuracy from each SVM is the mean accuracy from all SVMs trained for each pairwise comparison. Out of sample cross-validation is provided indirectly by comparison of the components that were consistent contributors to accurate classification for each of the four syndromes *versus* controls.

Next we investigated the relationship between the FTLD-associated metabolome and survival. Survival analysis was performed with cox proportional hazards regression. Only components selected by sequential feature selection in all disease *versus* control SVMs were used as predictor variables. Age, gender and FTLD-group were entered as covariates. SVM analyses were performed using LIBSVM in MATLAB R2018b (MathWorks).[22] Other statistical tests used MATLAB R2018b (Mathworks, USA).

## Results

Table 1 summarises the clinical groups. There were significant differences between FTLD (all diseases combined) and control samples in forty-nine out of 842 metabolites detected (two sample *t*-test, FDR p <0.01). The statistical significance of each metabolite is plotted against fold-change in Figure 2A. These metabolites did not cluster in one pathway but were distributed across a wide range of metabolic pathways. These included sixteen amino acid, seven energy and carbohydrate, three cofactor and vitamin, sixteen lipid, three nucleotide and four xenobiotic pathways (Table 2). Twenty-six of these metabolites remained significant with family wise error correction (Bonferroni corrected p<0.01), distributed across a wide range of pathways.

**Table 1:** Demographic and clinical summary metrics of study participants. ACER: Addenbrookes Cognitive Examination – Revised, CBI: Cambridge Behavioural Inventory – Revised, PSP-RS (Progressive Supranuclear Palsy Rating Scale). P values are result of ANOVA across rows for all FTLD subgroups and Controls (where applicable), except %Male where a Chi squared test was used: ns = p>0.05.

|  | **FTLD**<br>**(all subgroups)** | **bvFTD** | **nvPPA** | **PSP** | **CBS** | **Control** | **p value**<br>**(FTLD vs**<br>**Control)** |
| --- | --- | --- | --- | --- | --- | --- | --- |
| **Number** | 134 | 30 | 26 | 45 | 33 | 32 |  |
| **Mean age at blood test**<br>**(SD)** | 70.36<br>(8.21) | 64.51<br>(7.17) | 72.00<br>(7.66) | 72.9<br>(8.06) | 70.91<br>(7.04) | 68.73<br>(9.03) | ns |
| **% Male** | 50 | 50 | 38 | 62 | 55 | 56 | ns |
| **Estimated onset to**<br>**phlebotomy (years) (SD)** | 4.86<br>(2.86) | 5.56<br>(2.91) | 4.59<br>(2.1) | 4.71<br>(3.12) | 4.68<br>(2.84) |  | ns |
| **Diagnosis to phlebotomy**<br>**(years) (SD)** | 1.52<br>(1.73) | 2.0<br>(2.11) | 1.64<br>(1.46) | 1.04<br>(1.39) | 1.68<br>(1.82) |  | ns |
| **Mean ACE-R (<100)**<br>**(SD)** | 62<br>(27) | 52<br>(30) | 61<br>(29) | 72<br>(22) | 61<br>(29) |  | 0.009 |
| **Mean CBI (<180)**<br>**(SD)** | 61<br>(28) | 83<br>(26) | 39<br>(32) | 56<br>(31) | 66<br>(34) |  | <0.001 |
| **Mean PSP-RS (<100)**<br>**(SD)** | - | - | - | 43 (15) | - |  | NA |

**Table 2:** Table of metabolites that were significantly different in FTLD syndromes combined, compared to healthy controls. P value columns show the p value for a generalised linear model between FTLD and controls with age and sex as covariates. P-values in the uncorrected column in bold indicate survival after Bonferroni correction (equivalent to uncorrected p<1.33e^-5^)

| Metabolite Name | Subpathway | Superpathway | Fold change | P value (FDR) | P value (uncorr) |
| --- | --- | --- | --- | --- | --- |
| guanidinoacetate | Creatine | Amino Acid | 0.73 | 5.73E-06 | **1.14E-07** |
| beta-citrylglutamate | Glutamate | Amino Acid | 1.47 | 6.33E-05 | **1.93E-06** |
| 1-pyrroline-5-carboxylate | Glutamate | Amino Acid | 1.60 | 3.14E-03 | 1.58E-04 |
| 2-aminobutyrate | Glutathione | Amino Acid | 0.77 | 2.19E-03 | 9.57E-05 |
| sarcosine | Glycine/Serine/ Threonine | Amino Acid | 0.76 | 9.13E-08 | **8.48E-10** |
| 2-methylserine | Glycine/Serine/ Threonine | Amino Acid | 0.51 | 7.02E-11 | **9.31E-14** |
| N-acetylmethionine | Methionine/Cysteine/Sam/ Taurine | Amino Acid | 1.22 | 9.61E-03 | 6.25E-04 |
| alpha-ketobutyrate | Methionine/Cysteine/Sam/ Taurine | Amino Acid | 0.38 | 2.53E-10 | **1.01E-12** |
| hypotaurine | Methionine/Cysteine/Sam/ Taurine | Amino Acid | 1.98 | 1.03E-06 | **1.51E-08** |
| taurine | Methionine/Cysteine/Sam/ Taurine | Amino Acid | 1.63 | 7.68E-08 | **5.14E-10** |
| spermidine | Polyamine | Amino Acid | 3.42 | 1.12E-04 | **3.69E-06** |
| 5-methylthioadenosine (MTA) | Polyamine | Amino Acid | 1.24 | 5.34E-03 | 2.91E-04 |
| tryptophan betaine | Tryptophan | Amino Acid | 0.45 | 5.90E-03 | 3.50E-04 |
| serotonin | Tryptophan | Amino Acid | 10.71 | 1.22E-05 | **3.23E-07** |
| homoarginine | Urea Cycle; Arginine/ Proline | Amino Acid | 0.79 | 5.78E-03 | 3.30E-04 |
| pro-hydroxy-pro | Urea Cycle; Arginine/ Proline | Amino Acid | 1.40 | 4.77E-03 | 2.53E-04 |
| N-acetylneuraminate | Aminosugar | Carbohydrate | 1.47 | 1.14E-05 | **2.83E-07** |
| N-acetylglucosaminylasparagine | Aminosugar | Carbohydrate | 1.74 | 2.10E-03 | 8.93E-05 |
| maltotetraose | Glycogen | Carbohydrate | 16.02 | 1.14E-05 | **2.58E-07** |
| maltotriose | Glycogen | Carbohydrate | 10.87 | 7.68E-08 | **6.11E-10** |
| maltose | Glycogen | Carbohydrate | 3.08 | 1.50E-06 | **2.58E-08** |
| pyruvate | Glycolysis/Gluconeogenesis/ Pyruvate | Carbohydrate | 0.64 | 3.13E-03 | 1.45E-04 |
| nicotinamide | Nicotinate/ Nicotinamide | Cofactors/ Vitamins | 2.18 | 1.50E-06 | **2.48E-08** |
| adenosine 5'-diphosphoribose (ADP-ribose) | Nicotinate/ Nicotinamide | Cofactors/ Vitamins | 5.19 | 2.70E-05 | **7.52E-07** |
| flavin adenine dinucleotide (FAD) | Riboflavin | Cofactors/ Vitamins | 1.30 | 2.10E-03 | 8.87E-05 |
| succinate | TCA Cycle | Energy | 0.79 | 8.55E-03 | 5.33E-04 |
| stearamide | Fatty Acid/Amide | Lipid | 0.72 | 3.41E-03 | 1.76E-04 |
| pristanate | Fatty Acid/Branched | Lipid | 0.63 | 8.63E-04 | 3.09E-05 |
| maleate | Fatty Acid/Dicarboxylate | Lipid | 0.55 | 1.14E-05 | **2.88E-07** |
| glycerol 3-phosphate | Glycerolipid | Lipid | 0.66 | 7.69E-07 | **1.02E-08** |
| 1-(1-enyl-palmitoyl)-GPE (P-16:0)* | Lysoplasmalogen | Lipid | 1.25 | 5.77E-03 | 3.21E-04 |
| heptanoate (7:0) | Medium Chain Fatty Acid | Lipid | 1.86 | 1.85E-06 | **3.43E-08** |
| 10-undecenoate (11:1n1) | Medium Chain Fatty Acid | Lipid | 0.63 | 6.67E-06 | **1.42E-07** |
| 1-palmitoleoylglycerol (16:1) | Monoacylglycerol | Lipid | 0.46 | 1.65E-03 | 6.35E-05 |
| 1-linoleoylglycerol (18:2) | Monoacylglycerol | Lipid | 0.59 | 4.87E-04 | 1.68E-05 |
| 1-stearoyl-2-oleoyl-GPS (18:0/18:1) | Phosphatidylserine (PS) | Lipid | 8.94 | 1.95E-09 | **1.03E-11** |
| 1-stearoyl-2-arachidonoyl-GPS (18:0/20:4) | Phosphatidylserine (PS) | Lipid | 7.47 | 3.82E-07 | **4.56E-09** |
| choline phosphate | Phospholipid | Lipid | 1.59 | 1.42E-07 | **1.50E-09** |
| phosphoethanolamine | Phospholipid | Lipid | 2.61 | 9.92E-11 | **2.63E-13** |
| sphinganine | Sphingolipid | Lipid | 1.53 | 3.14E-03 | 1.55E-04 |
| sphingosine | Sphingolipid | Lipid | 1.38 | 2.57E-03 | 1.16E-04 |
| lactosyl-N-behenoyl-sphingosine (18:1/22:0) | Sphingolipid | Lipid | 1.48 | 3.14E-03 | 1.50E-04 |
| N1-methylinosine | Purine/(Hypo)Xanthine/Inosine Containing | Nucleotide | 1.46 | 5.90E-03 | 3.52E-04 |
| dihydroorotate | Pyrimidine/Orotate Containing | Nucleotide | 0.55 | 1.80E-03 | 7.18E-05 |
| 2'-deoxyuridine | Pyrimidine/Uracil Containing | Nucleotide | 0.65 | 1.62E-03 | 6.02E-05 |
| benzoate | Benzoate | Xenobiotics | 0.73 | 1.12E-04 | **3.72E-06** |
| iminodiacetate (IDA) | Chemical | Xenobiotics | 1.19 | 3.13E-05 | **9.15E-07** |
| thioprolin | Chemical | Xenobiotics | 1.16 | 8.69E-03 | 5.53E-04 |
| 1-methylxanthine | Xanthine | Xenobiotics | 0.58 | 6.34E-03 | 3.87E-04 |

**Figure 2:**
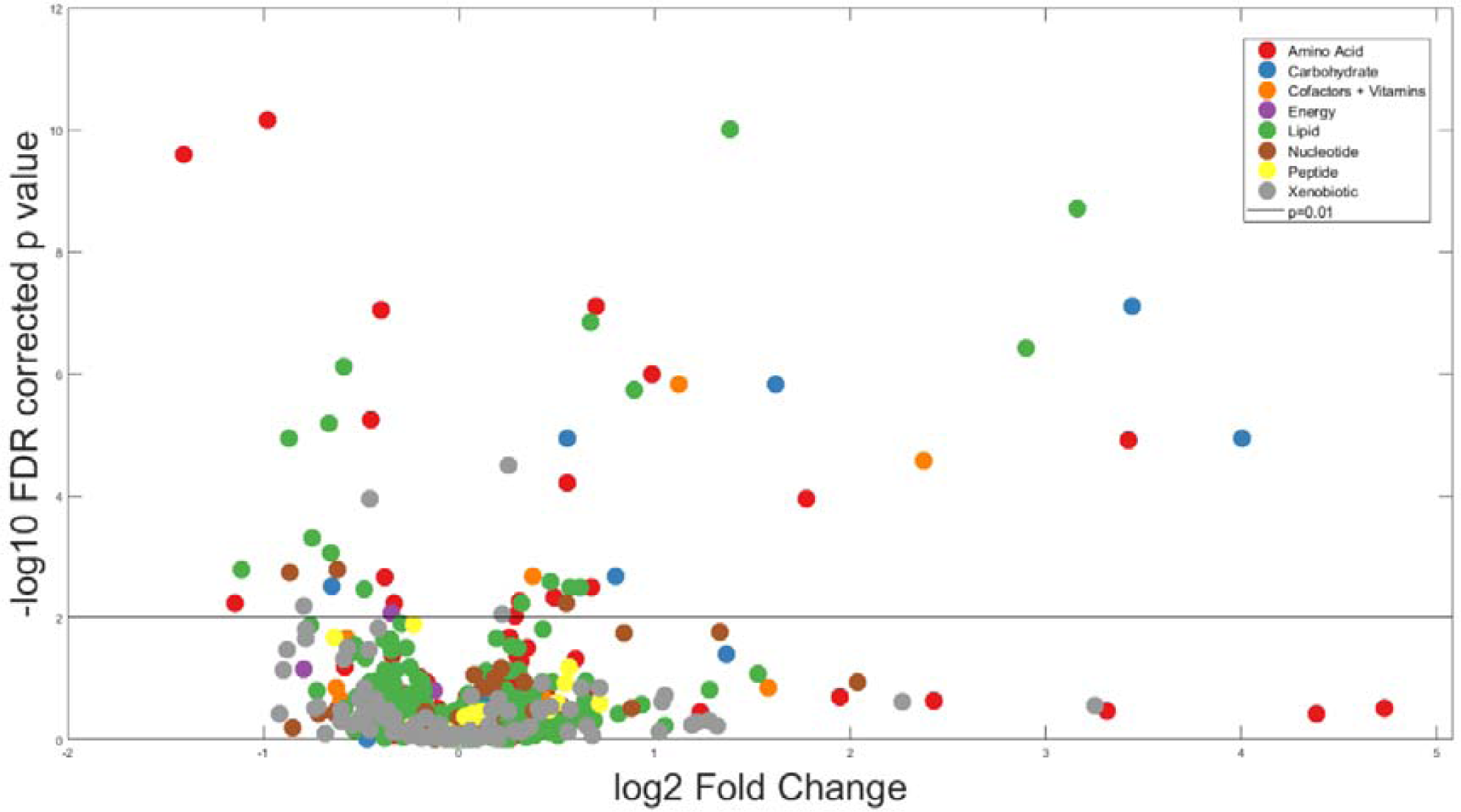
Metabolomic alterations in FTLD syndromes. Volcano plot of log-fold change in each metabolite for the contrast of FTLD vs control, and their significance (log-FDR corrected p-value). Metabolites are colour coded by superpathway. The horizontal line marks p=0.01 significance. The significant metabolites above this line, both increased and decreased, come from each the major metabolic pathways.

To assess differences in metabolic pathways, as opposed to individual metabolites, we compared the component loadings of principal component analyses on the metabolites in each pathway. Principle component analysis on each of 91 sub-pathways yielded 230 individual components. The component scores on twelve sub-pathways were significantly different between FTLD and controls (two sample *t*-test, FDR p <0.01). These included widespread changes in the metabolome including amino acid (creatine, glutamate, glycine, serine, threonine, methionine, cysteine, taurine, polyamine and tryptophan), carbohydrate (amino sugar and glycogen) and lipid (fatty acid, lysoplasmalogen, mevalonate, monoacylglycerol and phospholipid) pathways.

We then tested the efficacy of metabolomics as a diagnostic biomarker for FTLD (Table 3). Linear support vector machines with sequential feature selection using all 50 principle components from the global PCA as predictor variables accurately distinguished FTLD from controls (92.5%) and individual FTLD syndromes from controls (bvFTD 96.67% nfvPPA 88.08% PSP 95.78% CBS 95.16%). Accuracy was less among FTLD syndromes. BvFTD classification accuracy from nfvPPA (82.00%), PSP (81.33%) and CBS (83.33%) was better than PSP, CBS and nfvPPA. This was even lower in separating nfvPPA from PSP (79.52%) or CBS (0.72%) and PSP from CBS (78.6%).

**Table 3:** Matrix of average classification accuracy of the support vector machines’ classification between groups (percentage total correct classification). Groups were sized matched for each classifier (see methods). The diagonal values represent the classification accuracy for that disease group against all other groups combined. Classification accuracy is high in each FTLD syndrome compared with healthy controls, but lower when classifying between FTLD syndromes. [bvFTD=behavioural variant frontotemporal dementia, nfvPPA=non-fluent variant primary progressive aphasia, PSP=progressive supranuclear palsy Richardson’s syndrome, CBS=corticobasal syndrome].

|  | bvFTD | nfPPA | PSP | CBS | Control |
| --- | --- | --- | --- | --- | --- |
| bvFTD | 86 | 82 | 81 | 83 | 97 |
| nfPPA | 82 | 80 | 76 | 72 | 88 |
| PSP | 81 | 76 | 83 | 79 | 96 |
| CBS | 83 | 72 | 79 | 82 | 95 |
| Control | 97 | 88 | 96 | 95 | 93 |

Sequential feature selection, by removing components that did not contribute to SVM accuracy, identifies the components that best separated the two groups. Only 2 or 3 components were selected for each disease vs control classifier. One principle component was selected in every comparison between disease group and controls (component 3). From the between syndrome classifications, additional components were consistently selected (up to 6 in the bvFTD vs CBS comparison). For the nfvPPA vs CBS classifier accuracy no components were consistently selected.

Component 3, from the global PCA of all metabolite pathways, was selected by sequential feature selection in every disease vs control classifier. This means the metabolites contributing to this component were consistently important in separating disease groups from controls. All but two healthy controls positively loaded onto this component while the loadings in the FTLD syndromes varied (Figure 3A). Component 3 represented metabolites from a wide range of pathways (Figure 3B, full list of subpathway loadings in Appendix 2). Sub-pathways with high positive loading onto component 3 included phospholipid and other lipid pathways, haemoglobin and the carbohydrate glycogen metabolism pathway. Subpathways with high negative loading onto component 3 included certain fatty-acid pathways and amino acid pathways including leucine, valine, tryptophan, glycine, serine, threonine, methionine, cysteine and taurine metabolism.

**Figure 3A:**
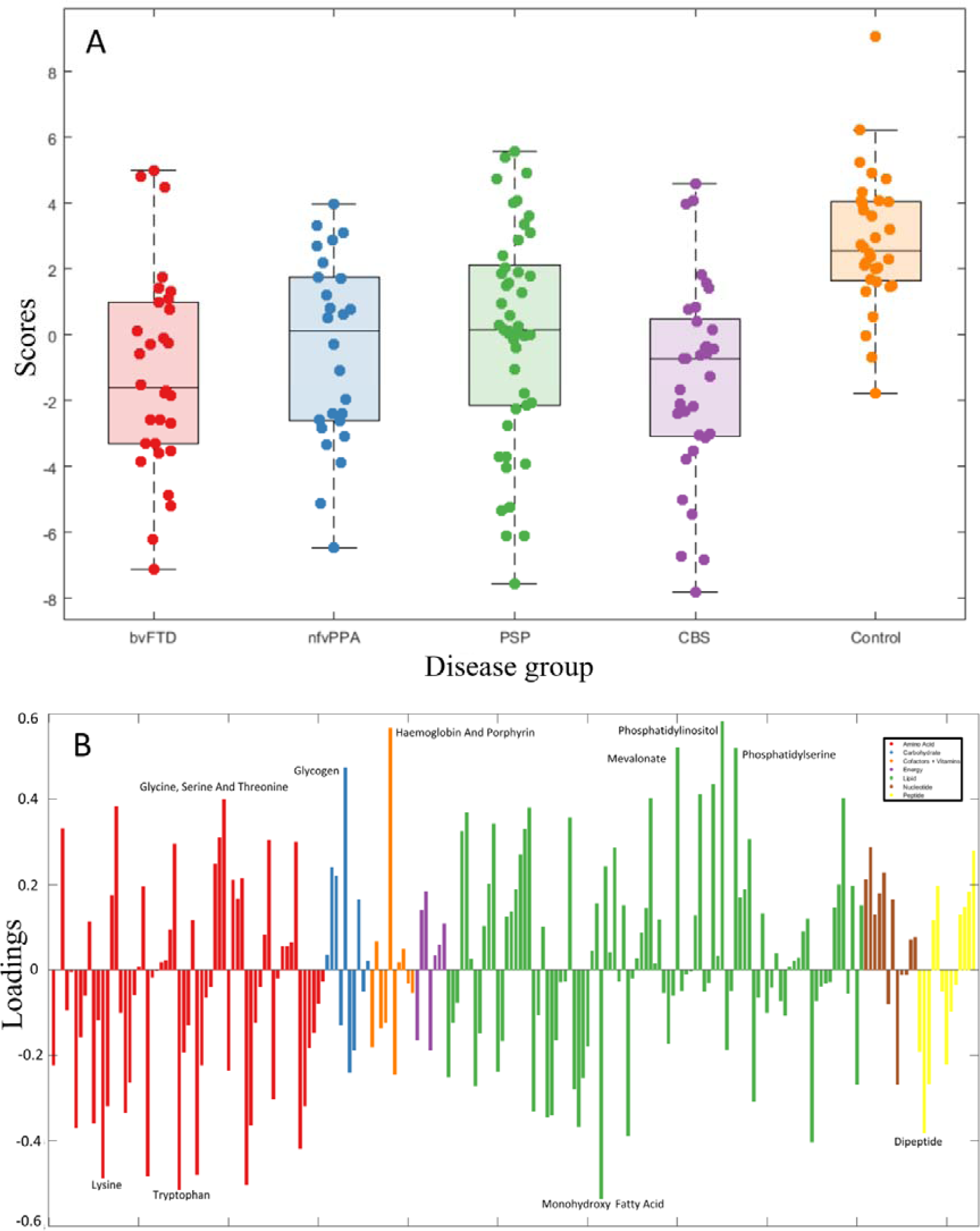
Individual loading onto component 3, by group. 3B. Subpathways loading on component 3.

We next tested component 3 as a prognostic biomarker (in patients only) using Cox proportional hazards regression using age, gender, disease groups and component 3 and days from blood test to death. The standardised individual participant loadings on component 3 were significantly associated with time to death (hazard ratio 0.74 (0.59-0.93), p=0.0018). To illustrate this effect, we plotted separately the patients with high (z score>1), medium (z score between 1 and -1) and low (z score<1) values on this component (Figure 4).

**Figure 4:**
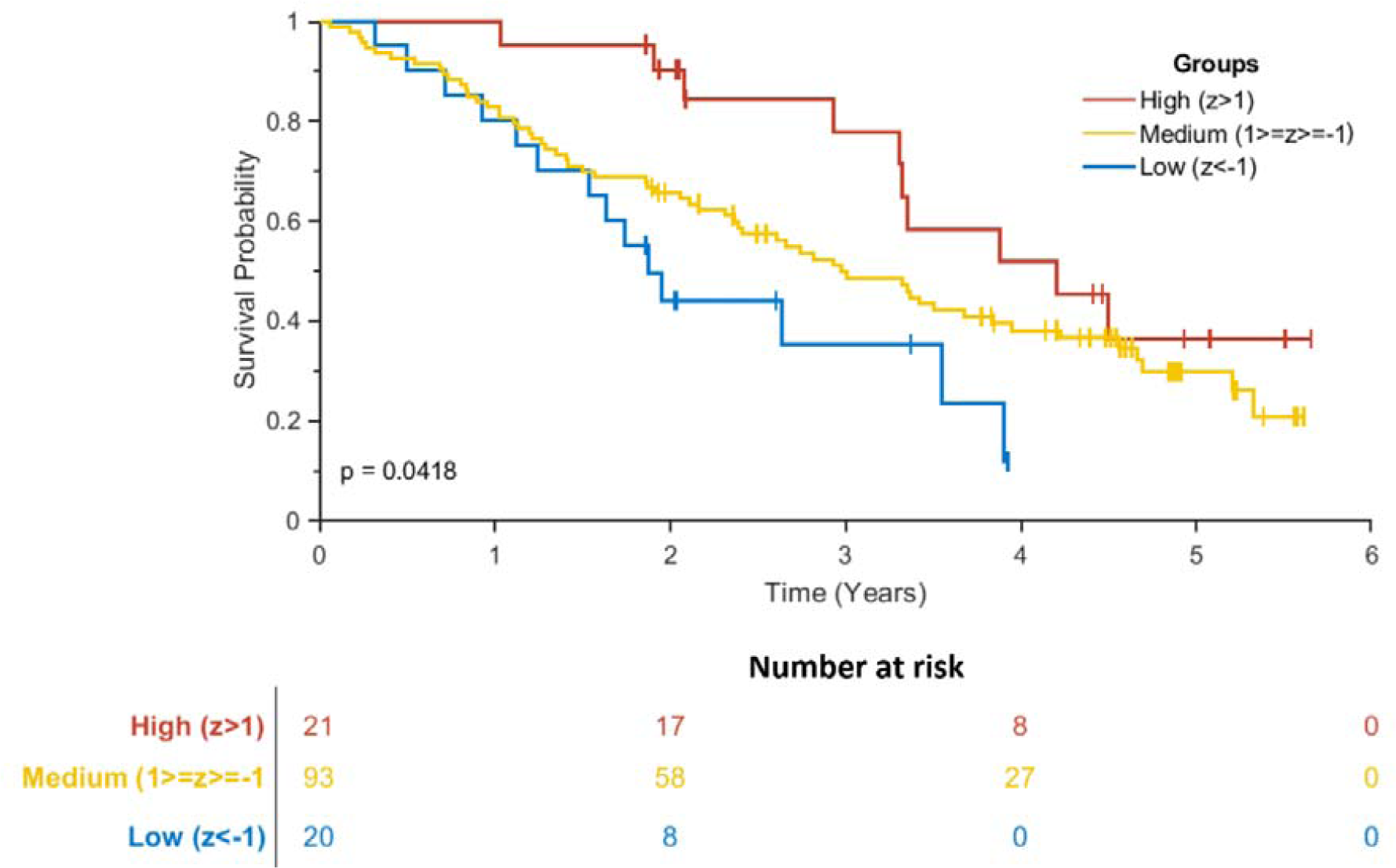
Kaplan-Meir Survival Curve of loadings on component 3. Patients were separated into three groups based on their loading onto component 3. High loading patients had a z score greater than 1, medium between 1 and -1 and low had a z score less than -1. There was a significant difference in survival curves between the three groups (log rank p=0.04). Graph generated using MatSurv (https://github.com/aebergl/MatSurv)

## Discussion

Our results show that multiple metabolic pathways are changed in patients with clinical syndromes associated with frontotemporal lobar degeneration. One profile of metabolic change (here identified as component 3) was consistently identified as feature of each disease *versus* controls, and the degree to which a patient expressed this metabolomic profile was correlated with subsequent survival. The metabolic changes in FTLD were not confined to a single pathway but were found across carbohydrates, lipids, amino acids, and peptide pathways. The identification of a blood-based metabolic index of FTLD and survival could assist differential diagnosis and clinical trial design, but we acknowledge that it is not known whether these abnormalities result from specific aetiopathogenic processes, or environmental sequelae of neurodegenerative disease. Replication in independent cohorts and the analysis of longitudinal change will also be important extensions of this work. In the following, we discuss the metabolic changes in turn, their potential utility for diagnosis and prognosis, and the study’s limitations.

Our first aim was to identify metabolic markers of FTLD. Several of the metabolite differences in FTLD implicate carbohydrate metabolism and energy pathways. Maltose and maltose metabolites, elevated in our FTLD groups, are primary disaccharides in the human diet. This result may be due to the altered dietary preferences, appetite, weight change and exercise associated with FTLD.[23–26] However, it may also be due to endogenous changes in energy metabolism and storage. Pyruvate and succinate, both key components of the TCA cycle, were low in FTLD despite the raised polysaccharides levels. Glycerol-3-phosphate, which has an important role in reoxidisation of NADH, was also low. These abnormalities reflect altered glucose uptake and metabolic dysfunction, which is of particular interest in view of *in vivo* PET imaging of FTLD where abnormal glucose metabolism often precedes neuronal loss and atrophy.[27–29]

The amino acid differences could also be attributed to defective energy metabolism. For example, glucogenic amino acid metabolites, including alpha-ketobutyrate, 2-methylserine and sarcosine were low in FTLD and in other neurodegenerative disease it has been suggested that abnormalities in these pathways represent an attempt to preserve or restore glycolysis.[30] Spermidine, elevated in FTLD, is a polyamine amino acid that promotes autophagy and has neuroprotective effects in rodent models.[31] The raised levels in FTLD might reflect a response to increased cell death that occurs in patients with neurodegenerative disease.[32] We found increased serotonin levels in FLTD (FC: 10.71, p<0.001). Central nervous serotonergic pathways are abnormal in FTLD [33] and serotonin reuptake inhibitors have been used as a symptomatic treatment in FTLD.[34] However, there is usually limited exchange of serotonin across the blood brain barrier, and the significance of this peripheral serotonin result is unclear for the central nervous system. Peripheral serotonin effects include glucose regulation via its action on pancreatic beta cells, hepatocytes and adipose tissue.[35] Abnormal peripheral serotonin levels in FTLD may therefore again relate to abnormal glucose regulation.

Lipid pathways were also abnormal in FTLD with alterations in several phospholipid, glycerolipid and sphingolipid metabolites. These are important components of cell membranes. Phospholipid pathway metabolites, including phosphatidylserines (FC7-8, p<0.001) and phosphoethanolamine (FC 2.61, p<0.001), showed the greatest differences in FTLD compared to controls. Our results contrast with a lipidomics study of bvFTD which found the same phospholipids were *reduced* in bvFTD. However, the apparent discrepancy could be explained by the differences in disease stages. Phospholipids are a major component of cell membranes and phosphatidylserine has been proposed as a pro-apoptotic marker in pre-clinical neuronal models of tauopathies.[36, 37] Sphingosine and its derivative sphingoamine, important components of sphingolipid metabolism, were also elevated in FTLD syndromes. Sphingosine derived lipids comprise up to one third of cell membranes and are highly prevalent in central nervous system white matter. Dysregulated sphingomyelin metabolism has been implicated in neurodegeneration due to Alzheimer’s disease [38] and have been suggested as a potential blood biomarker.[39]

Our second aim was to determine whether the metabolome could be used to classify patients by syndrome and provide proof-of-concept for a blood-based biomarker. Classification accuracy, using only the metabolite principle components, was high (88-97%) between each FTLD syndrome and controls. Sequential feature selection found that only a small subset of components was required, without loss of accuracy. Interestingly classification accuracy did not reflect the strength of the published clinicopathological correlations in each syndrome. Frontotemporal lobar degeneration syndromes are associated with different underlying pathologies, including FTLD-tau and FTLD-TDP43.[1] Each FTLD syndrome has a different clinicopathological accuracy; the clinical syndrome of PSP-Richardson’s syndrome is almost always caused by 4-repeat tau pathology [17] and had a classification accuracy of 95%. BvFTD, which can be caused by Tau-, TDP43-or FUS-pathology [15], still had a metabolomics accuracy of 96.5%. CBS has poor clinic-pathological correlation and may be associated with Corticobasal degeneration, Alzheimer’s Disease pathology, PSP or other pathology [40], but the syndrome still manifested a metabolomic classification accuracy of 95.6%. This would suggest some of the classifying features results are not neuropathologically specific but rather reflect generalised aspects of progressive neurodegeneration or the widespread physiological stresses that follow.

Classification accuracy was lower between the different FTLD syndromes. This is expected in view of the closely overlapping clinical features and underlying neuropathologies across FTLD syndromes. We suggest that the FTLD syndromes with the closest overlap in phenotype and pathology have the lowest classification accuracy. For example, nfvPPA can be the initial presenting syndrome of PSP-pathology or corticobasal degeneration, and nfvPPA can evolve towards a CBS-like phenotype, or CBS-NAV.[18, 41–43] PSP and CBS were weakly differentiated, and share many similar features in pathology and syndrome, as indicated by the nosological status of PSP-CBS and CBS-PSP.[17, 18]

Our third aim was to find a prognostic biomarker in FTLD. Component 3 was associated with survival (days to death), independent of disease group, age or gender. A range of metabolic pathways contributed to this component, including phospholipid, amino acid, carbohydrate and cofactor pathways. This suggests the metabolomics marker of mortality risk reflects a signature of underlying progressive neurodegeneration, as opposed to an isolated metabolic pathway alteration. We suggest that the component reflects both environmental and endogenous changes, but further studies are required to target the biochemicals comprising component 3. Despite the uncertainty over the causes of the metabolomic differences, our findings suggest that blood-based biomarkers have potential as diagnostic biomarkers. To confirm the role of metabolomics as a prognostic biomarker longitudinal measures are essential, and comparisons against other differential diagnostic groups such as Parkinson’s disease and non-degenerative causes of late-life behavioural change.

Our study has several limitations. Metabolomics can be highly sensitive to differences in sampling, storage and analysis. For practical reasons, and with a view to utility in healthcare settings, our samples were taken at variable times of day, and participants were not fasted. For ethical reasons, no medications were withheld or altered in participants for the purposes of the study. Some participants were taking levodopa or selective serotonin reuptake inhibitors for example. This might account for some of differences between disease groups and controls. However, to mitigate this risk, we removed metabolites and sub-pathways that have been associated with these medications in reference datasets. We also acknowledge that the Metabolon analysis pipeline cannot differentiate between optical isomers of a metabolite, which may have different physiological properties. Our sample size is modest, we restricted our classification sample sizes to prevent inequalities in the group sizes (which may otherwise bias a classifiers). Our sample was therefore limited by the prevalence of the least common disorder. Nonetheless, for small sizes, the k-fold cross-validation approach provides a minimally biased estimate of the potential accuracy of classification. For each disease group, we used within-sample cross validation, separating training and tests data on each iteration, but we have not replicated our findings in an independent cohort. Out-of-sample cross-validation was found across the four separate disease groups for component 3, which was most closely associated with survival. However, further work is required to replicate the findings in other disease-specific cohorts to confirm the utility of metabolomics as a diagnostic biomarker. In anticipation of clinical utility, we focussed on comparison and classification by syndrome. However, genetic FTD cohorts and retrospective analysis samples from people with post mortem diagnostic confirmation would enable the additional metabolomics analysis by pathology rather than syndrome.

In summary, our findings highlight the widespread metabolic changes in each of four major clinical disorders associated with frontotemporal lobar degeneration. We found that the metabolite profile can be used to classify between FTLD and healthy controls with high accuracy and relate to prognosis. Several metabolites show promise as diagnostic and prognostic biomarkers which if developed could enrich case identification in healthcare settings and in future clinical trials. Further work is required to replicate these findings and test their utility in differentiating between FTLD and pathologically distinct, but phenotypically similar syndromes.

## Data Availability

Anonymised data are available on reasonable request for academic purposes.

## Acknowledgements

We are grateful to Dr Matthew Davey, University of Cambridge for advice on statistical procedures and Dr Edward Karolyn from Metabolon Inc for his advice on the Metabolon analysis pipeline

## Compliance with ethical standards

### Ethical approval

This study received ethical approval from the Cambridge Central Research Ethics Committee (12/EE/0475 and 15/EE/0270) and was carried out in accordance with the ethical standards laid down in the 1964 Declaration of Helsinki and its later amendments.

### Informed consent

Participants with capacity gave written informed consent. The participation of those who lacked capacity was discussed with a consultee in accordance with the Mental Capacity Act (UK).

